# Susceptibility and infectiousness of children and adults with SARS-CoV-2 variant B.1.1.7 deduced from three daycare centre outbreaks and related household situations; Germany, 2021

**DOI:** 10.1101/2021.05.12.21256608

**Authors:** Anna Loenenbach, Inessa Markus, Ann-Sophie Lehfeld, Matthias an der Heiden, Walter Haas, Maya Kiegele, André Ponzi, Barbara Unger-Goldinger, Cornelius Weidenauer, Helen Schlosser, Alexander Beile, Udo Buchholz

**Author notes:** contributed equally (Anna Loenenbach, Inessa Markus). **Corresponding author:** Anna Loenenbach **Corresponding author email:**.

## Abstract

We investigated three SARS-CoV-2 variant B.1.1.7 kindergarten outbreaks and related household situations. Despite group cohorting, cases occurred in almost all groups, i.e. also among persons without close contact. Secondary attack rates (SAR) of children were similar to adults (day care: 23% vs. 30%; p=0.15; households: 32% vs. 39%; p=0.27), and also child-induced household outbreaks led to similar SAR compared to adults. With the advent of B.1.1.7, susceptibility and infectiousness of children and adults seem to converge.

## Introduction

Before the circulation of the SARS-CoV-2 “variants of concern” (VOC), children have been associated with lower susceptibility compared to adults (1-3). They were rarely identified as primary cases in household studies (4) and gave rise to a (secondary) attack rate ((S)AR) of 7.9% (95% confidence interval (CI), 1.7%-16.8%) (3). In German childcare centres, children have led to an average SAR of 1.7% and small outbreak sizes (average size: 3-4 cases) (5). For contact tracing purposes, Germany distinguished between high-risk contacts for a contact with a COVID-19 case within 1.5 metre distance for 15 minutes or longer and low-risk contacts for contacts for >1.5 metre distance and/or for less than 15 minutes. High-risk contacts were associated with a SAR of 5% (6) to 13% (7), while low-risk contacts were associated with a SAR of 0% (6) to 3% (7).

In December 2020, the SARS-CoV-2 “variant of concern” (VOC) B.1.1.7 (N501Y.V1) began to circulate in Germany. As of 3 March 2021, approximately 40% of all randomly selected swabs of all COVID-19-cases were tested positive for VOC B.1.1.7 (8). While several studies have presented evidence for the increased transmissibility of B.1.1.7 (9-11), the susceptibility and infectiousness of children of preschool age compared to previously circulating strains was still unknown.

In January/February 2021, three B.1.1.7 kindergarten outbreaks occurred almost simultaneously in a district of one local health authority (LHA). Together with the related household situations these outbreaks gave us the opportunity to assess (1) if high-risk contact definitions still hold, (2) if there are indications of increased susceptibility of preschoolers in comparison to adults, and (3) the infectiousness of preschoolers, particularly in comparison to adults in household settings.

We investigated the outbreaks and collected information regarding structural, organizational and infection control measures in the facilities. We assessed SAR within the kindergartens and associated households with particular focus on type of contact and age.

### Description of the outbreaks and analysis

All three kindergartens had the following measures in place: elaborate hygiene and infection control plans; cohort grouping with reduced number of children and designated staff, and access to separate bathroom areas; playing outside only within groups in assigned playground areas; staff wearing masks at least outside of the group rooms; prohibition for parents to enter the building and obligation to wear masks during childrens’ handover; digital meetings with staff/parents, if possible. We visited the facilities and conducted interviews with kindergarten management and primary and/or first secondary cases. Comprehensive testing was offered to all persons at the facilities. All contact persons (CP) were followed-up daily for symptoms by the LHA. Any person becoming symptomatic was tested. We defined a close contact as an encounter of ≥15 minutes within a distance of <1.5 m. All other persons in the kindergarten were defined as non-close contacts.

The overall SAR in the three kindergartens were 37%, 27% and 17% (Table 1). In all three outbreaks the likely primary cases (PC) were adults, none of the reported symptoms at the day of symptom onset was respiratory, but rather general, such as fatigue, headache, back pain or exhaustion. In all three kindergartens, 80% to 88% of secondary cases occurred within an interval of 7 days (Table 1). Eleven (92%) of all twelve cohorted groups had secondary cases. SAR for close CP were 53%, 33% and 22%, and among non-close contacts 32%, 26% and 6%. SAR for children were 31%, 27% and 17%, and among adults 53%, 28% and 17% (Table 1).

**Table 1:** Secondary attack rates (SAR) of SARS-CoV-2 B.1.1.7 in sub-groups, by type of contact and by age in three kindergarten outbreaks; Germany, 2021

|  |  | Kindergarten 1 |  |  | Kindergarten 2 |  |  | Kindergarten 3 |  |  |
| --- | --- | --- | --- | --- | --- | --- | --- | --- | --- | --- |
|  |  | n/N | SAR in % | 95% CI) | n/N | SAR in % | 95% CI | n/N | SAR in % | 95% CI |
| Overall SAR |  | 25/68 | 37 | 26-49 | 12/44 | 27 | 16-42 | 10/59 | 17 | 9-28 |
| 7 days clustering * |  | 22/25 | 88% |  | 10/12 | 83% |  | 8/10 | 80% |  |
| Group |  |  |  |  |  |  |  |  |  |  |
|  | Group 1 (with PC) | 8/15 | 53 | 30-75 | 3/9 | 33 | 12-65 | 6/18 | 33 | 16-56 |
|  | Group 2 | 7/11 | 64 | 35-85 | 6/15 | 40 | 20-64 | 3/20 | 15 | 5-36 |
|  | Group 3 | 2/9 | 22 | 6-55 | 1/7 | 14 | 3-51 | NA |  |  |
|  | Group 4 | 5/16 | 31 | 14-56 | NA |  |  | NA |  |  |
|  | Daycare | 2/12 | 17 | 5-45 | 0/7 | 0 | 0-35 | 1/15 | 7 | 1-30 |
|  | Staff | 1/5 | 20 | 4-62 | 2/6 | 33 | 10-70 | 0/6 | 0 | 0-39 |
| Type of contact |  |  |  |  |  |  |  |  |  |  |
|  | close contact | 8/15 | 53 | 30-75 | 3/9 | 33 | 12-65 | 9/41 | 22 | 12-37 |
|  | no close contact | 17/53 | 32 | 21-45 | 9/35 | 26 | 14-42 | 1/18 | 6 | 1-26 |
| Age |  |  |  |  |  |  |  |  |  |  |
|  | children | 15/49 | 31 | 20-45 | 7/26 | 27 | 14-46 | 6/36 | 17 | 8-32 |
|  | adults | 10/19 | 53 | 32-73 | 5/18 | 28 | 12-51 | 4/23 | 17 | 7-37 |

SAR: secondary attack rates; CI: confidence interval; PC: primary case
Facility contact persons testing negative or in the absence of a test (kindergarten 1: n=13; kindergarten 2: n=3; kindergarten 3: n=15) without symptoms were counted as non-case.
\* Clustering of secondary cases within in an interval of 7 days. For asymptomatic persons ((kindergarten 1: n=7; kindergarten 2: n=5; kindergarten 3: n=2) no day of symptom onset was known; instead the day of testing was assigned.

Households were not tested systematically. However, we offered CP a test shortly after the PC tested positive. We contacted households to monitor symptom onsets and tested symptomatic persons. Household CP testing negative or, in the absence of a test (19/92; 21%) without symptoms, were counted as non-cases. After exclusion of single households and households with another possible PC, we included 38 households with 92 CP. Pooled household SAR was 37% (95% CI: 28-47; Table 2). If the household CP was a child (32%), SAR was lower but not significantly different compared to adult household CP (39%). SAR in households were (also not significantly) higher when the PC was a child (39% vs 33% for adult PC).

**Table 2:** Secondary household attack rates (SAR) among household contacts of primary cases stemming from three kindergarten outbreaks; Germany, 2021.

| Household situation |  | n/N | SAR in % (95% CI) | RR (95% CI) |
| --- | --- | --- | --- | --- |
| Overall |  | 34/92 | 37 (28-47) |  |
| if household contact person is a child |  | 9/28 | 32 (18-51) | 0.82 (0.44-1.52) |
| if household contact person is an adult |  | 25/64 | 39 (28-51) | reference |
| if PC is a child (n=22) |  | 23/59 | 39 (28-52) | 1.17 (0.66-2.09) |
|  | if household contact person is a child | 4/15 | 27 (11-52) |  |
|  | if household contact person is an adult | 19/44 | 43 (30-58) |  |
| if PC is an adult (n=16) |  | 11/33 | 33 (20-50) | reference |
|  | if household contact person is a child | 5/13 | 38 (18-64) |  |
|  | if household contact person is an adult | 6/20 | 30 (15-52) |  |

SAR: secondary attack rates; CI: confidence interval; RR: relative risk; PC: primary case

## Discussion

These simultaneously occurring kindergarten outbreaks provided a good opportunity to revisit definitions of “closeness”. At least in the first two outbreaks, CP not fulfilling the existing definition of a “close contact” had a substantially higher SAR than previously reported. Moreover, in all three outbreaks, we documented case-clustering within approximately a week’s time, which is compatible with a one- or two-day exposure for the entire kindergarten. Remarkably, transmission occurred in similar proportions to children and adults alike. It occurred to eleven out of twelve groups, including the rather separated daycare groups. We have gained a good overview of attitudes and practices of educators and the adherence to hygiene and infection control measures in the kindergartens. For example, it enabled us to group the two kindergarten groups of kindergarten 3 together as “close contact” because they took their meals together. Furthermore, having visited the kindergartens one-by-one we observed that room sizes and ventilation differed significantly and could have contributed as a possible transmission factor. Kindergarten 1 had the smallest volume per person, kindergarten 2 had a ventilation system and two groups in kindergarten 3 were spread out over two floors. In addition, the third kindergarten was shut down entirely more quickly than the other kindergartens which might explain its smaller outbreak size.

With the exception of the first kindergarten, susceptibility of children, as illustrated by the SAR in the other kindergartens and the household outbreaks, was similar as in adults and substantially higher than described for the pre-VOC strains. Household SAR among children and adults in this study (32% and 39%, respectively) were substantially higher compared to non-VOC household SAR as assessed in a meta-analysis (17% and 28%, respectively) (3). Anecdotal evidence on B.1.1.7-outbreaks in United States daycare centers indicate that SAR may occur at surprisingly high rates (12, 13). Data from the United Kingdom on AR among B.1.1.7 “contacts” (not further defined) stratified in 10-year age groups indicate a similar relative SAR increase among most age groups (14). However, reported SAR among 0-9-year-olds are still lower compared to adults (9% vs. approximately 20%).

Important questions of this investigation were, if children – after having been infected in kindergarten - lead to further infections in their household, and if so to what extent do children infect adults in their households. The above mentioned meta-analysis found a (non-significantly) lower SAR when children were the PC (7.9%; 95% CI, 1.7%-16.8%) compared to SAR when adults were the household PC (15.2%; 95% CI, 6.2%-27.4%) (3). In our study, we found corresponding SAR of 39% (95% CI: 28-52%) for child PCs and 33% for adult PCs (95% CI: 20-50%).

We acknowledge the following limitations: First, because not everyone was tested, we may have underestimated the true SAR. However, symptom monitoring during quarantine was in place and would have led to immediate testing. Second, in households, asymptomatic persons may be the true PC, and cases outside the family may have infected a family member independently from the assumed household’s PC. Even if these possibilities may be true in single households it is unlikely to distort gravely the overall picture.

In summary, our investigation confirms increased transmissibility of B.1.1.7. In addition, the data presented suggest that both susceptibility and infectiousness of children 1-5-years-old is substantially higher compared to the pre-VOC era, and may be converging to those among adults. To prevent individual daycare center outbreaks, or at least limit outbreak size, measures need to be revisited, including non-pharmaceutical measures, early closure should be considered when cases are occurring and vaccination of educators should be promoted.

## Supporting information

Ethics_Statement

## Data Availability

No external dataset was used.

## Conflict of interest

None.

## Funding statement

The work was conducted by persons employed by the government or the local health department. We do not have to declare any other source of funding.

## Notes

### Competing Interest Statement

The authors have declared no competing interest.

### Funding Statement

No external funding was received, that should be reported.

### Author Declarations

The German Protection against Infection Act and the law on the duties of the Robert Koch Institute (RKI) allowed the implementation of this outbreak investigation without seeking further institutional ethical review (Infektionsschutzgesetz, IfSG, BGBl. I S. 1045 https://www.rki.de/DE/Content/Infekt/IfSG/ifsg_node.html; https://www.gesetze-im-internet.de/ifsg/index.html). Informed consent was obtained from participants. As deputy head of the Department for Infectious Disease Epidemiology at the RKI, the national Public Health Institute of Germany, certified that: - She is the competent authority for assessing whether outbreak investigations and/or research require review by an institutional ethics committee or if the German Protection against Infection Act (Infektionsschutzgesetz, IfSG, BGBl. I S. 1045) allows investigation and/or research without additional institutional review. - The outbreak investigation presented by Loenenbach & Markus et al. in 'Susceptibility and infectiousness of children and adults with SARS-CoV-2 variant B.1.1.7 deduced from three daycare centre outbreaks and related household situations; Germany, 2021' was conducted as part of the official tasks of the local public health authorities of the respective district, supported by the RKI upon official request in accordance to paragraph 4 of the German Protection against Infection Act. Therefore, this investigation was exempt from additional institutional review. We are happy to provide a signed letter with this declaration. All necessary patient/participant consent has been obtained and the appropriate institutional forms have been archived.

