## Supplementary material for "Susceptibility and infectiousness of children and adults with SARS-CoV-2 variant B.1.1.7 deduced from three daycare centre outbreaks and related household situations; Germany, 2021": Ethics_Statement

ROBERT KOCH INSTITUT

Robert Koch Institute | Nordufer 20 | 13353 Berlin

Department for Infectious Disease Epidemiology

To whom it may concern

Subject: Susceptibility and infectiousness of children and adults with SARS-CoV-2 variant B.1.1.7 deduced from three daycare centre outbreaks and related household situations; Germany, 2021

10.05.2021

As head of the Department for Infectious Disease Epidemiology at the Robert Koch Institute (RKI), the national Public Health Institute of Germany, I hereby certify that: I represent the competent authority for assessing whether outbreak investigations and/or research require review by an institutional ethics committee or if the German Protection against Infection Act (Infektionsschutzgesetz, IfSG, BGBl. I S. 1045) allows investigation and/or research without additional institutional review.

The outbreak investigation presented by Loenenbach & Markus et al. in "**Susceptibility and infectiousness of children and adults with SARS-CoV-2 variant B.1.1.7 deduced from three daycare centre outbreaks and related household situations; Germany, 2021**" was conducted as part of the official tasks of the local public health authorities of the respective district, supported by the RKI upon official request in accordance to §4 of the German Protection against Infection Act. Therefore, this investigation was exempt from additional institutional review.

Sincerely,

Dr. Ute Rexroth

Deputy Head of Department for Infectious Disease Epidemiology

Robert Koch Institute

Robert Koch Institute

www.rki.de

Reporting/
Processing by: A. Loenenbach, I. Markus

Extension: -2065/-3792
E-Mail:
 /


Address:
Nordufer 20
13353 Berlin
Federal Republic of Germany

The Robert Koch Institute is a federal institute within the portfolio of the Federal Ministry of Health
